# Adherence to the Mediterranean Dietary Approaches to Stop Hypertension Intervention for Neurodegenerative Delay (MIND) Diet and Trajectories of Depressive Symptomatology in Youth

**DOI:** 10.1101/2024.12.08.24318688

**Authors:** Yiwei Pu, Hangyu Tan, Runqi Huang, Wenchong Du, Qiang Luo, Tai Ren, Fei Li

## Abstract

**Background:** The rising prevalence of youth depression underscores the need to identify modifiable factors for prevention and intervention. This study aims to investigate the protective role of Mediterranean-DASH Intervention for Neurodegenerative Delay (MIND) diet on depressive symptoms in adolescents.

**Methods:** Participants were identified from the Adolescent Brain Cognitive Development study. Adherence to the MIND diet was measured by the Child Nutrition Assessment or the Block Kids Food Screener. Depressive symptoms were measured annually using the Child Behavior Checklist’s depression subscale. We utilized regression analyses and cross-lagged panel modeling (CLPM) to examine longitudinal associations. Additional analyses adjusted for polygenic risk scores for depression, and changes in Body Mass Index (BMI) and waist-to-height ratio.

**Results:** Of the 8,459 children (52.3% male; mean age 10.9 [SD, 0.6] years), 2,338 (27.6%) demonstrated high MIND diet adherence, while 2,120 (25.1%) showed low adherence. High adherence was prospectively associated with reduced depressive symptoms (adjusted β, -0.64, 95% CI, -0.73 to -0.55) and 46% lower odds of clinically relevant depression (adjusted odds ratio, 0.54, 95% CI, 0.39 to 0.75) at two-year follow-up. CLPM analyses showed significant cross-lag paths from MIND diet scores to less depressive symptoms across three time points. These associations persisted independently of changes in BMI and waist-to-height ratios, and were not significantly moderated by genetic predisposition to depression.

**Conclusions:** Higher adherence to the MIND dietary pattern was longitudinally associated with decreased risk of depressive symptoms in adolescents. Promoting MIND diet may represent a promising strategy for depression prevention in adolescent populations.

## Introduction

Depression is a leading cause of worldwide disability, exhibiting a rising prevalence and high recurrence rates^[1]^. Adolescent-onset depression are commonly accompanied by pronounced functional impairments, including additional psychiatric, somatic comorbidities, and more likely recurrent episodes^[2]^. Recent epidemiological data suggest a concerning upward trend in youth depression rates, necessitating identification of modifiable risk factors for prevention and intervention^[3]^.

Dietary patterns have emerged as critical modulators of depression risk, with robust evidence supporting the protective effects of the Mediterranean diet^[4-6]^. However, this dietary pattern’s strict requirements, particularly its restricted food choices, present significant adherence challenges for children and adolescents^[7]^. While the Mediterranean diet’s antidepressant effects may operate through anti-inflammatory pathways and optimization of oxidative stress, alternative dietary patterns with broader food selections might achieve comparable benefits^[8]^. The Mediterranean-DASH Intervention for Neurodegenerative Delay (MIND) diet represents a more flexible approach, combining key elements from the Mediterranean and Dietary Approaches to Stop Hypertension (DASH) diets while expanding food choices, designed to emphasize foods potentially protective against cognitive decline^[9]^. The MIND diet expands protein options beyond the Mediterranean diet’s emphasis on seafood, while preserving key neuroprotective nutrients such as flavonoids, folate, and omega-3 fatty acids^[9]^. Studies examining the MIND diet’s impact on depression in adults have yielded mixed results, potentially due to cohort heterogeneity and varying depression etiologies across different age groups^[10, 11]^. Notably, despite adolescence being a critical period for both dietary habit formation and depression onset, the potential protective effects of the MIND diet in this age group remain unexplored.

This study leveraged the data from the Adolescent Brain Cognitive Development (ABCD) study, a prospective cohort of over 10,000 children across the United States. We investigated the temporal relationship between MIND diet adherence and depressive symptoms during adolescent development.

## Methods

### Study population

This study utilized data from Release 5.1 of the ABCD Study, a large-scale longitudinal cohort comprising approximately 20% of the U.S. population within the target age demographic^[12, 13]^. The ABCD Study enrolled a total of 11,876 children between 9 and 11 years of age through school systems across 21 research sites in the United States between 2016 and 2018, with ongoing annual follow-ups. Our analytical sample included participants with complete data for both the Child Behavior Checklist (CBCL) and Food Frequency Questionnaire across the initial three years of follow-up. All participants and their guardians provided informed consent.

### Measures

#### MIND diet

Dietary assessment utilized two instruments across the study period. During the first-year follow-up, parents completed the Child Nutrition Assessment (CNA), an adapted version of the MIND diet questionnaire. The original MIND diet questionnaire evaluates consumption frequency across 15 food categories: whole grains, vegetables, leafy greens, berries, nuts, poultry, legumes, fish, wine, olive oil, fast food, fried foods, pastries, confectionery, butter, and cheese. The CNA adaptation employed in the ABCD study excluded the wine category, resulting in 14 food categories (Table S1). Responses were binary-coded (yes=1, no=0) and summed to established a score indicating adherence to MIND diet (range, 0 to 14). Adherence to MIND diet was trichotomized as low (0-6), moderate (7-9), and high (10-14) using established cut-points derived from tertiles in population^[14]^.

For the second- and third-year follow-up, dietary habits were collected using the Block Kids Food Screener (BKFS), reported by either children or their parents. Given the BKFS structure, the MIND score was modified to include 10 food categories, omitting 5 categories (Table S2). Responses were binary-coded (yes=1, no=0) and summed to represent the adherence to MIND diet (range, 0 to 10). The MIND diet scores across various time points showed moderate and comparable correlation coefficients (first- and second-year, r=0.22; second- and third-year, r=0.34; Table S3). For the BKFS-derived MIND score, we also trichotomized the study population by the tertiles as low (0-2), moderate (3-4), and high (5-6) at each follow-up.

### Depressive symptom

Depressive symptoms were evaluated annually through parental reports using the CBCL. The CBCL is a widely used measure of youth emotional and behavioral problems including 120 questions rating on three-point Likert scale: 0 (not true), 1 (somewhat/sometimes true), and 2 (very/often true). A composite score for depression was calculated by summing all relevant questions, according to the manual of the CBCL (Diagnostic and Statistical Manual of Mental Disorders [Fifth Edition]-oriented scales). Higher scores indicate more depressive symptoms^[15]^. In addition, the age- and sex-adjusted T-score no less than 70 was corresponding to higher than the 98^th^ percentile in the general population, thus defined as depression. We also examined a similar CBCL-derived score of withdrawn/depressed symptom, to assess the robustness of our results.

### Polygenic Risk Score for Depression

Genotype data from the ABCD Study were sourced from saliva or blood specimens using the Affymetrix NIDA Smoke Screen Array^[16]^. A subset of 5,807 individuals of European descent were selected based on genetic lineage. Quality control and imputation were conducted using PLINK v1.90^[17]^, Michigan Imputation Server^[18]^, and Eagle v2.4, resulting in 4,673 samples for analysis. To construct PRS for depressive symptoms, we incorporated data from the genome-wide analyses of depressive symptoms (n = 161,460) of European ancestry^[19]^, using a continuous shrinkage with a global shrinkage prior of 0.01^[20]^. The PRS were trichotomized into tertiles, designated as low, moderate risk, and high risk.

### Covariates

We consider the following covariates as confounders: age at assessment, sex (male, or female), race/ethnicity (White, Black, Hispanic, Asian, or other), family annual income (<$35,000, $35,000∼$75,000, $75,000∼$100,000, or ≥$100,000), Body Mass Index (BMI, kg/m^2^) at assessment, parental average education year, and pubertal score at assessment. The pubertal score was the average of self- and parent-reported score on a scale from 1 (prepuberty) to 5 (post puberty). BMI was converted into sex and age-specific z-scores in accordance with Centers for Disease Control and Prevention (CDC) growth curves^[21]^. For variables with substantial missing data at certain follow-ups (specifically, BMI at third follow-up), values were carried forward from the most recent previous assessment. The waist-to-height ratio has been proposed as a simple and effective adiposity metric to assess central obesity widely applied to pediatric populations^[22]^.

### Statistical analysis

Chi-square test and t-test were performed to compare demographic characteristics among children at different level of adhering to the MIND diet. To investigate the correlation between MIND score and depression, we applied mixed-effect regressions to account for the nested structure of family and site. Linear regression was applied for continuous outcomes, including depressive symptom scores, while logistic regression was applied for bivariate outcomes, including depression. To examine the temporal relationship between MIND and depressive symptoms, we applied the cross-lagged panel model (CLPM), a type of structural equation model for analyzing the unidirectional or reciprocal relationships between variables measured at multiple time points. The goodness-of-fit was measured and compared by log-likelihood, chi-square, Comparative Fit Index, Tucker-Lewis Index, and Root Mean Square Error of Approximation. To explore the potential influence of overweight on the relationship between MIND diet scores and depressive symptoms, we conducted stratified sensitivity analyses based on whether BMI exceeded the 85th percentile (p85) and the additionally adjusting for waist-to-height ratio at assignment. In sensitivity analyses, imputation of missing BMI of the third-year follow-up was conducted by the average of the second and fourth-year follow-ups, followed by the conduct of sensitivity analyses.

We performed all analyses using R software version 4.2.2. CLPM model was performed by package lavaan (version 0.6-17). Mixed models were fitted with the package lme4 (version 1.1-35.3). Missing values were imputed by chained equations with package mice (version 3.16.0).

## Results

A total of 8,459 children had complete data of both the CBCL and dietary assessment questionnaires across three annual follow-ups. The mean (SD) age of participants was 10.9 (0.6) years at first follow-up, 12.0 (0.7) years at second follow-up, and 12.9 (0.6) years at third follow-up. Participants were categorized based on their MIND diet scores at the first follow-up. MIND diet adherence scores were significantly higher among female participants and those of Hispanic ethnicity (Table 1). Family income appeared to be higher among the moderate adherence group. Throughout the follow-up period, obesity measures were comparable among the three groups. The pubertal scores were comparable in the second-year follow-up, though a higher level in the high adherence group was observed at the first and third follow-ups.

**Table 1.**
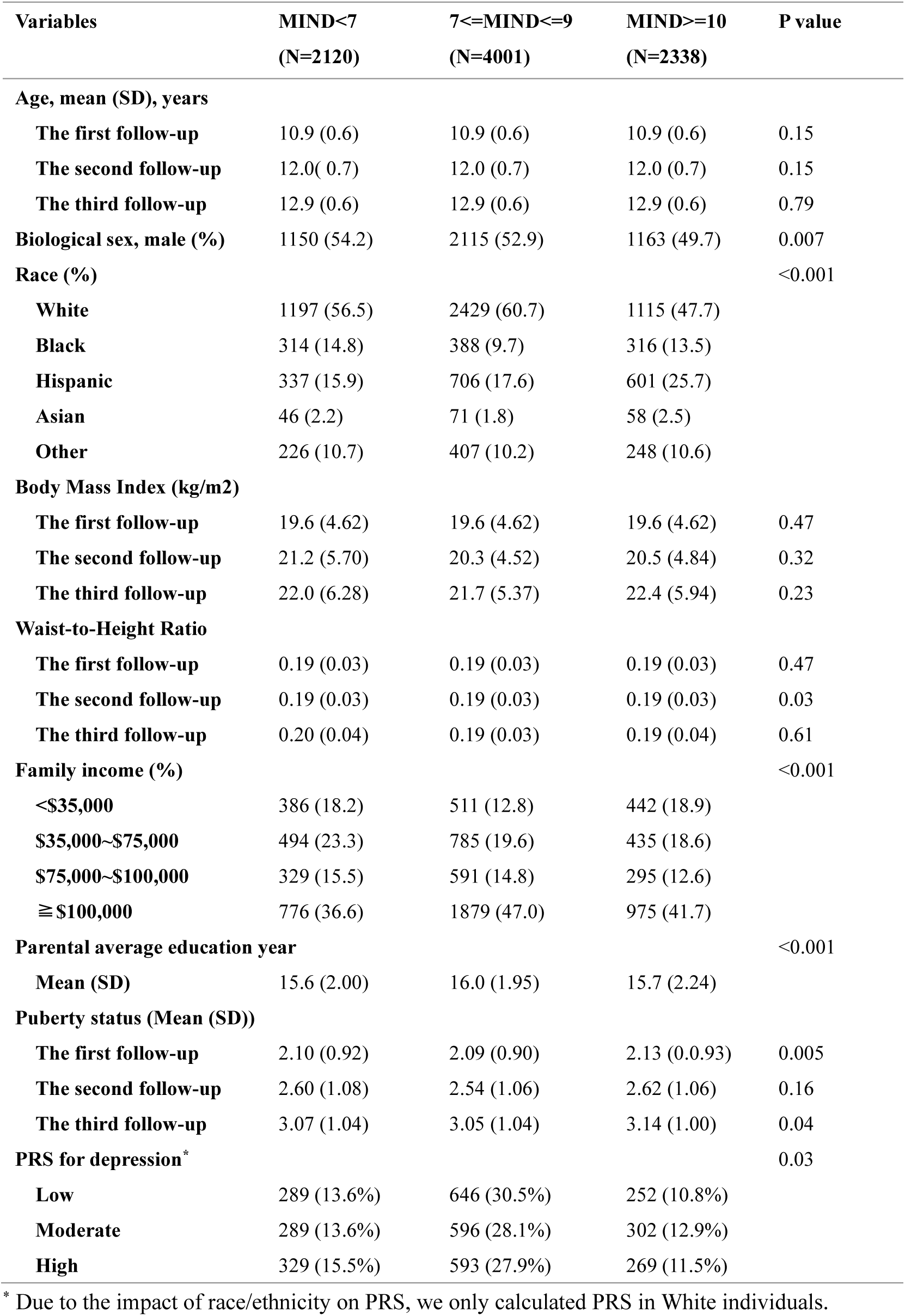
Demographics of the study population, categorized by their adherence to MIND diet. MIND, the mediterranean dietary approaches to stop hypertension intervention for neurodegenerative delay; PRS, polygenic risk score. The aggregate percentile distribution did not reach 100% owing to the presence of missing observations. Proportion of missing values: age at first-year follow-up, N=1; age at second-year follow-up, N=1; age at third-year follow-up, N=1; BMI at first-year follow-up, N=75; BMI at second-year follow-up, N=1249; BMI at third-year follow-up, N=6763; Waist-to-Height Ratio at first-year follow-up, N=68; Waist-to-Height Ratio at second-year follow-up, N=1277; Waist-to-Height Ratio at third-year follow-up, N=6771; family income at recruitment, N=561; Parental average education year, N=1485; puberty status at first-year follow-up, N=193; puberty status at second-year follow-up, N=399; puberty status at third-year follow-up, N=467; PRS for depression, N=4894.

| Variables | MIND<7<br>(N=2120) | 7<=MIND<=9<br>(N=4001) | MIND>=10<br>(N=2338) | P value |
| --- | --- | --- | --- | --- |
| **Age, mean (SD), years** |  |  |  |  |
| The first follow-up | 10.9 (0.6) | 10.9 (0.6) | 10.9 (0.6) | 0.15 |
| The second follow-up | 12.0 (0.7) | 12.0 (0.7) | 12.0 (0.7) | 0.15 |
| The third follow-up | 12.9 (0.6) | 12.9 (0.6) | 12.9 (0.6) | 0.79 |
| Biological sex, male (%) | 1150 (54.2) | 2115 (52.9) | 1163 (49.7) | 0.007 |
| Race (%) |  |  |  | <0.001 |
| White | 1197 (56.5) | 2429 (60.7) | 1115 (47.7) |  |
| Black | 314 (14.8) | 388 (9.7) | 316 (13.5) |  |
| Hispanic | 337 (15.9) | 706 (17.6) | 601 (25.7) |  |
| Asian | 46 (2.2) | 71 (1.8) | 58 (2.5) |  |
| Other | 226 (10.7) | 407 (10.2) | 248 (10.6) |  |
| **Body Mass Index (kg/m2)** |  |  |  |  |
| The first follow-up | 19.6 (4.62) | 19.6 (4.62) | 19.6 (4.62) | 0.47 |
| The second follow-up | 21.2 (5.70) | 20.3 (4.52) | 20.5 (4.84) | 0.32 |
| The third follow-up | 22.0 (6.28) | 21.7 (5.37) | 22.4 (5.94) | 0.23 |
| **Waist-to-Height Ratio** |  |  |  |  |
| The first follow-up | 0.19 (0.03) | 0.19 (0.03) | 0.19 (0.03) | 0.47 |
| The second follow-up | 0.19 (0.03) | 0.19 (0.03) | 0.19 (0.03) | 0.03 |
| The third follow-up | 0.20 (0.04) | 0.19 (0.03) | 0.19 (0.04) | 0.61 |
| Family income (%) |  |  |  | <0.001 |
| <\$35,000 | 386 (18.2) | 511 (12.8) | 442 (18.9) | |
| \$35,000~\$75,000 | 494 (23.3) | 785 (19.6) | 435 (18.6) | |
| \$75,000~\$100,000 | 329 (15.5) | 591 (14.8) | 295 (12.6) | |
| ≥\$100,000 | 776 (36.6) | 1879 (47.0) | 975 (41.7) | |
| Parental average education year |  |  |  | <0.001 |
| Mean (SD) | 15.6 (2.00) | 16.0 (1.95) | 15.7 (2.24) |  |
| **Puberty status (Mean (SD))** |  |  |  |  |
| The first follow-up | 2.10 (0.92) | 2.09 (0.90) | 2.13 (0.93) | 0.005 |
| The second follow-up | 2.60 (1.08) | 2.54 (1.06) | 2.62 (1.06) | 0.16 |
| The third follow-up | 3.07 (1.04) | 3.05 (1.04) | 3.14 (1.00) | 0.04 |
| PRS for depression* |  |  |  | 0.03 |
| Low | 289 (13.6%) | 646 (30.5%) | 252 (10.8%) |  |
| Moderate | 289 (13.6%) | 596 (28.1%) | 302 (12.9%) |  |
| High | 329 (15.5%) | 593 (27.9%) | 269 (11.5%) |  |

\* Due to the impact of race/ethnicity on PRS, we only calculated PRS in White individuals.

The mean depressive score in the low MIND diet adherence group was 1.87 (SD, 2.49) at the first-year follow-up, compared to 1.30 (SD, 2.03) in the moderate group and 0.94 (SD, 1.75) in the high adherence group. Using T-score ≥ 70 to define depression, the rates of depression were 5.4%, 2.8%, and 2.1% in the low, moderate, and high adherence group, respectively (Table S4). The high MIND diet adherence group was associated with a 0.74-point lower depressive score compared to the low adherence group, adjusting for age, sex, race, family income, parental education level, puberty status, and BMI (adjusted β, -0.74, 95% CI, -0.84 to -0.65; Figure 1). Likewise, the group with high adherence to the MIND diet exhibited a 60% reduction in the odds of depression compared to low adherence (adjusted OR, 0.40, 95% CI, 0.27 to 0.58; Figure 2). The moderate adherence group exhibited consistent and less prominent associations. Consistent cross-sectional associations were observed when utilizing the withdraw depression scale as the outcome measure (Table S8-9).

**Figure 1.**
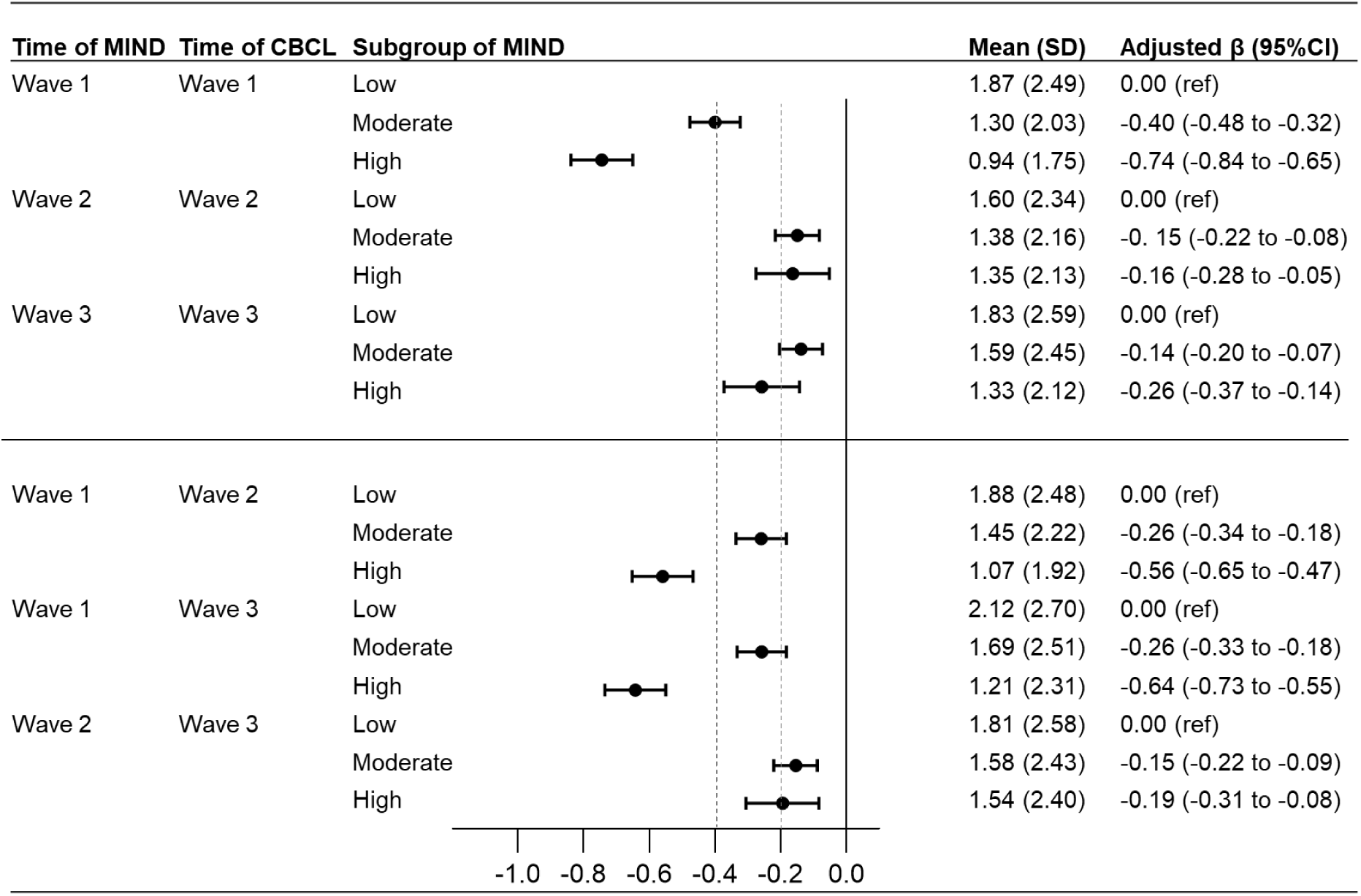
Cross-sectional and longitudinal associations between adherence to MIND diet and depression symptom score. All analyses were adjusted for age at assessment, sex, race/ethnicity, family annual income, Body Mass Index at assessment, parental average education year, and pubertal score at assessment. MIND, the mediterranean dietary approaches to stop hypertension intervention for neurodegenerative delay; PRS, polygenic risk score; CBCL, the Child Behavior Checklist.

**Figure 2.**
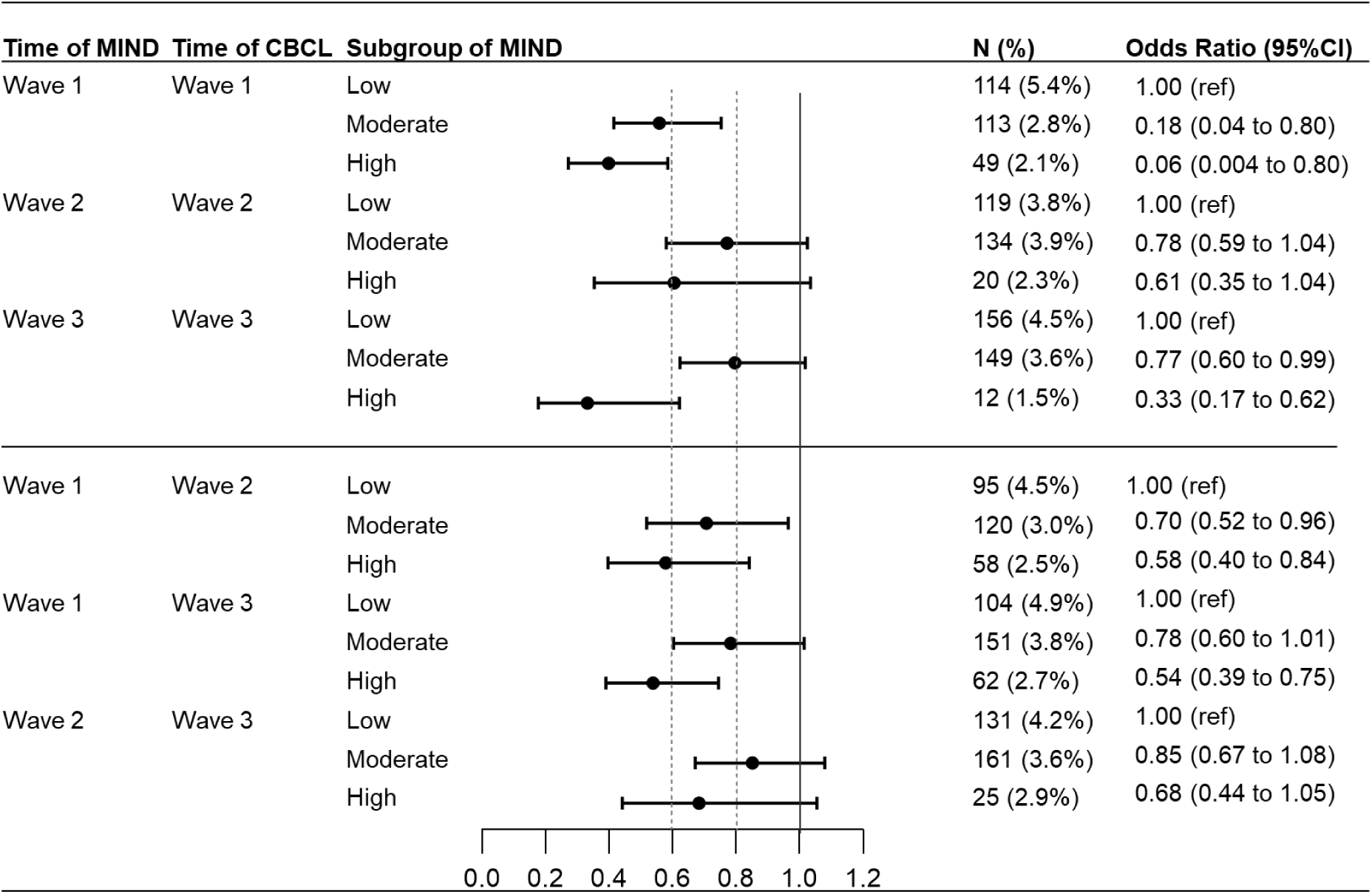
Cross-sectional and longitudinal associations between adherence to MIND diet and clinical depression. All analyses were adjusted for age at assessment, sex, race/ethnicity, family annual income, Body Mass Index at assessment, parental average education year, and pubertal score at assessment. MIND, the mediterranean dietary approaches to stop hypertension intervention for neurodegenerative delay; PRS, polygenic risk score; CBCL, the Child Behavior Checklist.

Longitudinally, high MIND diet adherence at the first follow-up was predictive of lower depressive scores at both second and third follow-ups, compared to the low adherence group (adjusted β for the second follow-up, -0.56, 95% CI, -0.65 to -0.47; adjusted β for the third follow-up, -0.64, 95% CI, -0.73 to -0.55; Figure 1). Likewise, the high MIND diet adherence group was associated with a lower odd of depression (Figure 2). The results were robust when additionally adjusting for the depressive scores at the first follow-up. Similar results were found between MIND score at the second follow-up and depressive score at the third follow-up (Figure 1, 2). Additionally adjusting for the depression symptom at the time of MIND yielded consistent results (Table S10). Similar longitudinal associations were consistently observed when using withdraw depression scale as outcome (Table S8-9) To further understand the role of BMI and waist-to-height ratio, the analyses stratified by BMI exceeding the p85 and with additional adjustment for waist-to-height ratio yielded consistent cross-sectional and longitudinal results (Table S5-7).

To determine the temporal relationship between MIND diet and depression score, we applied CLPM and found that high MIND diet adherence was longitudinally associated with lower depression score (Figure 3). Fit indices revealed that the model was a good fit to the observed data (Table S11). The auto-regressive coefficients of MIND diet and depression score were significant (all P < 0.001). We observed significant prospective paths from MIND diet at wave 1 to depression score at wave 2 and from MIND diet at wave2 to depression score at wave 3. A significant path from depression score to MIND diet was only observed from wave 2 to wave 3.

**Figure 3.**
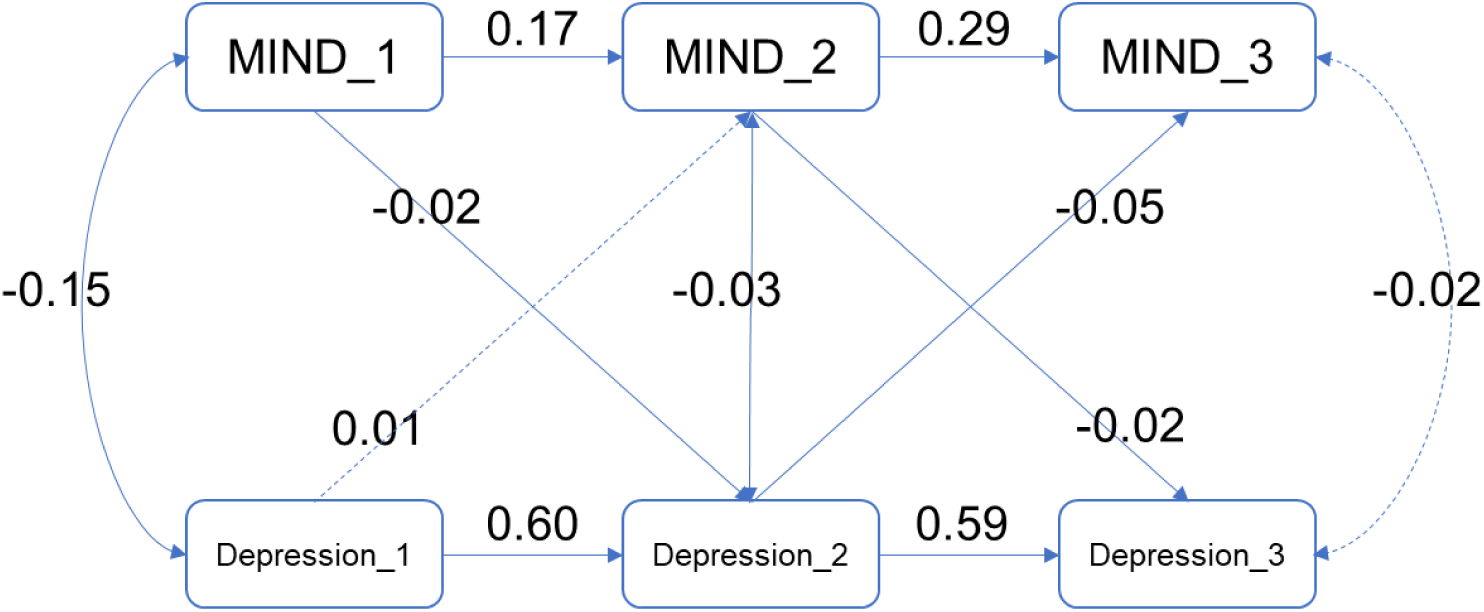
CLPM of MIND and depression symptom score. All analyses were adjusted for age at assessment, sex, race/ethnicity, family annual income, Body Mass Index at assessment, parental average education year, and pubertal score at assessment. CLPM, cross-lagged panel model; MIND, the mediterranean dietary approaches to stop hypertension intervention for neurodegenerative delay.

To investigate the role of genetic predisposition to depression, we included 3565 White children in analyses of PRS for depression symptoms. Categorizing the children by the tertiles of PRS for depression symptoms, we observed generally consistent cross-sectional and longitudinal association between adherence to MIND diet and lower depressive symptoms across PRS groups (Table S12). The risk estimates exhibited variations across comparisons, indicating limited evidence of moderation. Moreover, additionally adjusting for PRS for depression symptoms in the CLPM resulted in similar risk estimates of cross-lag paths (Figure S1).

## Discussion

In this large prospective cohort of adolescents, we found a robust longitudinal association of adherence to MIND diet with lower depressive symptoms and lower likelihood of having clinically significant depression at 1- and 2-year follow-up. MIND diet adherence demonstrated a dose-dependent relationship with depressive symptoms, where higher adherence levels predicted progressively lower symptom severity at follow-up assessments. These associations persisted independently of changes in BMI and waist-to-height ratios, and were not significantly moderated by genetic predisposition to depression.

Prior research on MIND diet and depression has focused primarily on adults. Only one study included participants under age 18 (n=19), but did not report outcomes specific to this adolescent subgroup^[23]^. In adults, one randomized trial showed that the MIND diet improved depressive symptoms among obese or overweight women with polycystic ovary syndrome^[24]^. Previous cohort studies have yielded inconsistent results, likely due to methodological limitations including cross-sectional designs and heterogeneous study populations spanning different age groups and body mass indices ^[24-26]^ ^[11, 27-29]^. Adolescence represents a critical period for both dietary habit formation and depression onset, yet research has not examined the potential protective effects of the MIND diet during this developmental stage. To our knowledge, we provide the first empirical evidence suggesting a potential protective effect of the MIND diet against depression in adolescents using a longitudinal design.

The MIND diet incorporates key components of the Mediterranean diet, which has been shown to reduce depressive symptoms in both adolescents and adults^[6]^. Both diets emphasize high intake of plants and complete protein, which provides elevated levels of polyphenols and polyunsaturated fatty acids - compounds that may improve depression through suppressing inflammatory^[30]^. Our finding aligns with established research showing that healthier dietary patterns correlate with decreased depressive symptoms. The MIND diet offers greater adherence potential for children and adolescents compared to the Mediterranean diet. For example, the MIND diet places a greater emphasis on the consumption of green leafy vegetables and berries, which are rich in fiber and thus can produce a higher amount of prebiotics^[31]^. These characteristics make the MIND diet particularly promising for enhancing mental health outcomes, including depression reduction, among children and adolescents^[32]^.

MIND diet is effective in weight control^[33]^. Previous studies suggested that adherence to the MIND diet may improve cognitive outcomes by reversing obesity-related brain alterations^[34]^. However, our study showed a significant association with depressive symptoms even adjusting for BMI or waist-to-height ratio at each follow-up, suggesting that obesity-related factors play a limited mediating role in the diet’s effects on depressive symptoms.

Genetic factors play a significant role in the development of depression among adolescents^[35]^. In investigating the role of genetic predisposition of depressive symptoms, we found limited evidence that the PRS moderated the association between MIND diet adherence and depressive symptoms. This suggests that the MIND diet’s protective effects may operate independently of genetic predisposition, highlighting its potential as a universally applicable modifiable factor for depression prevention.

Our study possesses several strengths. First, our longitudinal design and comprehensive series of analyses yielded robust results, enhancing the reliability of our findings. Second, this study is the first to investigate the protective role of the MIND diet against depressive symptoms specifically in adolescents. Third, the racial diversity of the ABCD study sample increases the generalizability of our findings across different populations.

Our findings should be interpreted with caution due to some limitations. First, the MIND diet assessments at waves 2 and 3 utilized questionnaires that were not specifically designed for MIND diet evaluation and lacked certain components. However, the questionnaires captured key dietary elements, and we demonstrated significant correlations between wave 1 (using standard MIND diet items) and wave 2 scores, aligning with previous research. Second, the absence of biological assessment data precluded investigation of potential underlying mechanisms, such as inflammatory markers. Future studies should examine these biological pathways to elucidate the mechanisms linking MIND diet adherence to depressive symptoms.

## Conclusion

Adhering to MIND diet was associated with less depressive symptoms risk over 2 years in children and adolescents. Our findings suggest that promoting MIND diet adherence may represent a promising strategy for depression prevention in adolescent populations.

## Data Availability

All data produced are available online at https://abcdstudy.org.

https://osf.io/cxjbf/?view_only=e54fb63f4aea46d58cba974aac4d6ae7

## Acknowledgements

Data used in the preparation of this article were obtained from the Adolescent Brain Cognitive Development^SM^ (ABCD) Study (https://abcdstudy.org), held in the NIMH Data Archive (NDA). This is a multisite, longitudinal study designed to recruit more than 10,000 children aged 9–10 and follow them over 10 years into early adulthood. The ABCD Study® is supported by the National Institutes of Health and additional federal partners under award numbers U01DA041048, U01DA050989, U01DA051016, U01DA041022, U01DA051018, U01DA051037, U01DA050987, U01DA041174, U01DA041106, U01DA041117, U01DA041028, U01DA041134, U01DA050988, U01DA051039, U01DA041156, U01DA041025, U01DA041120, U01DA051038, U01DA041148, U01DA041093, U01DA041089, U24DA041123, U24DA041147. A full list of supporters is available at https://abcdstudy.org/federal-partners.html. A listing of participating sites and a complete listing of the study investigators can be found at https://abcdstudy.org/consortium_members/. ABCD consortium investigators designed and implemented the study and/or provided data but did not necessarily participate in the analysis or writing of this report. This manuscript reflects the views of the authors and may not reflect the opinions or views of the NIH or ABCD consortium investigators. The ABCD data repository grows and changes over time. The ABCD data used in this report came from ABCD Dataset Data Release 5.1. An NDA study has been created for the data used in this report under the doi: 10.15154/d21c-1r22.

## Author Contributions

Dr Li had full access to all of the data in the study and take responsibility for the integrity of the data and the accuracy of the data analysis.

Concept and design: Fei Li, Tai Ren.

Acquisition, analysis, or interpretation of data: All authors.

Drafting of the manuscript: Yiwei Pu, Tai Ren, Hangyu Tan.

Critical review of the manuscript for important intellectual content: All authors.

Statistical analysis: Yiwei Pu, Tai Ren and Runqi Huang.

Obtained funding: Fei Li, Tai Ren.

Administrative, technical, or material support: All authors.

Supervision: Fei Li, Tai Ren.

## Availability of data and materials

Data are publicly released on an annual basis through the National Institute of Mental Health (NIMH) data archive (NDA, https://nda.nih.gov/abcd). The ABCD study are openly available to qualified researchers for free. Access can be requested at https://nda.nih.gov/abcd/request-access. The data that support the findings of this study are openly available in the ABCD Dataset Data Release 5.1. An NDA study has been created for the data used in this report and code for the replication of analyses conducted in the manuscript can be retrieved under the doi: 10.15154/d21c-1r22.

## Ethics approval and consent to participate

In the ABCD study, all procedures were approved by a central Institutional Review Board (IRB) at the University of California, San Diego, and in some cases by individual site IRBs (e.g., Washington University in St. Louis) (https://www.sciencedirect.com/science/article/pii/S1878929317300622). Parents or guardians provided written informed consent after the procedures had been fully explained and children assented before participation in the study.

