## Supplementary material for "Adherence to the Mediterranean Dietary Approaches to Stop Hypertension Intervention for Neurodegenerative Delay (MIND) Diet and Trajectories of Depressive Symptomatology in Youth": supplymentary

**Table of Contents**

Table S1. MIND Diet Components at First Year: Servings and Scores 2
Table S2. MIND Diet Components at Second and Third Year: Servings and Scores 3
Table S3. The correlation coefficients of the MIND diet scores across various time points 4
Table S4. the MIND Diet and depressive symptom T-score 5
Table S5. Cross-sectional and longitudinal associations between adherence to MIND diet and depression symptom score additionally adjusting for waist-to-height ratio at the assessment 6
Table S6. Cross-sectional and longitudinal associations between adherence to MIND diet and clinical depression additionally adjusting for waist-to-height ratio at the assessment 7
Table S7. Cross-sectional and longitudinal associations between adherence to MIND diet and depression symptom score, stratified based on whether BMI exceeds the 85th percentile 8
Table S8. Cross-sectional and longitudinal associations between adherence to MIND diet and withdraw depression score 9
Table S9. Cross-sectional and longitudinal associations between adherence to MIND diet and withdraw depression 10
Table S10. Longitudinal associations between adherence to MIND diet and depression symptom score, additionally adjusting for the depression symptom score at the assessment for MIND diet 11
Table S11. The predictive analysis between MIND and clinical depression additionally adjusting for whether clinical depression at the time for MIND diet 12
Table S11. CLPM model fit indices across waves 13
Table S12. Cross-sectional and longitudinal associations between adherence to MIND diet and depression symptom score additionally adjusting for PRS of depression symptoms. 14
Figure S1. CLPM of MIND and depression symptom score additionally adjusting for PRS. 15

**Table S1. MIND Diet Components at First Year: Servings and Scores**

| Diet component | Point awarded for adherence | | "Think about what your child eats in a typical week, during this past year since we last saw you. In a typical week, does your child eat____?" |
|---|---|---|---|
| | 0 | 1 | |
| **Whole grains (servings/day)** | ≤ 2 | ≥ 3 | Whole grains 3 or more times per day |
| **Green leafy vegetables (servings/week)** | ≤ 5 | ≥ 6 | Green leafy vegetables 6 or more times per week |
| **Other vegetables (servings/day)** | 0 | ≥ 1 | Other vegetables 1 or more time per day |
| **Berries (servings/week)** | ≤ 1 | ≥ 2 | Berries 2 or more times per week |
| **Red Meat (servings/week)** | ≥ 4 | < 4 | Red meats and meat products less than 4 times per week |
| **Fish (servings/week)** | 0 | ≥ 1 | Fish 1 or more time per week |
| **Poultry (servings/week)** | ≤ 1 | ≥ 2 | Poultry 2 or more times per week |
| **Beans (servings/week)** | ≤ 3 | ≥ 4 | Beans 4 or more times per week |
| **Nuts (servings/week)** | ≤ 4 | ≥ 5 | Nuts 5 or more times per week |
| **Fast food (servings/week)** | ≥ 1 | < 1 | Fast food or fried food less than 1 time per week |
| **Olive Oil (primary)** | no | yes | Olive oil is used as the primary oil |
| **Butter (servings/day)** | ≥ 1 | < 1 | Butter or margarine is used less than 1 tablespoon per day |
| **Cheese (servings/week)** | ≥ 1 | < 1 | Cheese less than 1 time per week |
| **Pastries (servings/week)** | ≥ 5 | < 5 | Pastries or sweets less than 5 times per week |

Question on wine consumption was omitted by the ABCD Study, as the adolescents were under the legal drinking age (i.e., 21) in the United States.

MIND, the mediterranean dietary approaches to stop hypertension intervention for neurodegenerative delay

**Table S2. MIND Diet Components at Second and Third Year: Servings and Scores**

| **Diet component** | **Point awarded for adherence** | | **Previous weeks frequency** |
|---|---|---|---|
| | 0 | 1 | |
| **Whole grains (servings/day)** | ≤ 2 | ≥ 3 | Whole grains 3 or more times per day |
| **Green leafy vegetables (servings/week)** | ≤ 5 | ≥ 6 | Green leafy vegetables 6 or more times per week |
| **Other vegetables (servings/day)** | 0 | ≥ 1 | Other vegetables 1 or more time per day |
| **Berries (servings/week)** | ≤ 1 | ≥ 2 | Berries 2 or more times per week |
| **Red Meat (servings/week)** | ≥ 4 | < 4 | Red meats and meat products less than 4 times per week |
| **Fish (servings/week)** | 0 | ≥ 1 | Fish 1 or more time per week |
| **Poultry (servings/week)** | ≤ 1 | ≥ 2 | Poultry 2 or more times per week |
| **Beans (servings/week)** | ≤ 3 | ≥ 4 | Beans 4 or more times per week |
| **Cheese (servings/week)** | ≥ 1 | < 1 | Cheese less than 1 time per week |
| **Pastries (servings/week)** | ≥ 5 | < 5 | Pastries or sweets less than 5 times per week |

MIND, the mediterranean dietary approaches to stop hypertension intervention for neurodegenerative delay

**Table S3. The correlation coefficients of the MIND diet scores across various time points**

| | MIND_1 | MIND_2 | MIND_3 |
|---|---|---|---|
| **MIND_1** | 1.00 | 0.22*** | 0.18*** |
| **MIND_2** | 0.22*** | 1.00 | 0.34*** |
| **MIND_3** | 0.18*** | 0.34*** | 1.00 |

MIND, the mediterranean dietary approaches to stop hypertension intervention for neurodegenerative delay

**Table S4. the MIND Diet and depressive symptom T-score**

| Variables | MIND<7 (N=2120) | 7<=MIND<=9 (N=4001) | MIND>=10 (N=2338) | Overall (N=8459) | P value |
|---|---|---|---|---|---|
| **cbcl_scr_dsm5_depress_t_1** | | | | | |
| **<70** | 2006 (94.6%) | 3888 (97.2%) | 2289 (97.9%) | 8183 (96.7%) | <0.001 |
| **>=70** | 114 (5.4%) | 113 (2.8%) | 49 (2.1%) | 276 (3.3%) | |
| **cbcl_scr_dsm5_depress_t_2** | | | | | |
| **<70** | 2025 (95.5%) | 3881 (97.0%) | 2280 (97.5%) | 8186 (96.8%) | <0.001 |
| **>=70** | 95 (4.5%) | 120 (3.0%) | 58 (2.5%) | 273 (3.2%) | |
| **cbcl_scr_dsm5_depress_t_3** | | | | | |
| **<70** | 2016 (95.1%) | 3850 (96.2%) | 2276 (97.3%) | 8142 (96.3%) | <0.001 |
| **>=70** | 104 (4.9%) | 151 (3.8%) | 62 (2.7%) | 317 (3.7%) | |

MIND, the mediterranean dietary approaches to stop hypertension intervention for neurodegenerative delay.

**Table S5. Cross-sectional and longitudinal associations between adherence to MIND diet and depression symptom score additionally adjusting for waist-to-height ratio at the assessment**

| Time of MIND | Time of CBCL | Subgroup of MIND | Mean (SD) | Adjusted β (95%CI) |
|---|---|---|---|---|
| **Wave 1** | Wave 1 | low | 1.89 (2.58) | 0.00 (ref) |
| | | moderate | 1.31 (2.03) | -0.38 (-0.49, -0.26) |
| | | high | 0.93 (1.74) | -0.72 (-0.86, -0.58) |
| **Wave 2** | Wave 2 | low | 1.56 (2.27) | 0.00 (ref) |
| | | moderate | 1.35 (2.10) | -0.18 (-0.28, -0.08) |
| | | high | 1.41 (2.19) | -0.14 (-0.30, 0.02) |
| **Wave 3** | Wave 3 | low | 1.74 (2.37) | 0.00 (ref) |
| | | moderate | 1.65 (2.50) | -0.08 (-0.18, 0.02) |
| | | high | 1.24 (1.98) | -0.32 (-0.49, -0.14) |
| **Wave 1** | Wave 2 | low | 1.84 (2.43) | 0.00 (ref) |
| | | moderate | 1.44 (2.14) | -0.25 (-0.37, -0.14) |
| | | high | 1.07 (1.92) | -0.54 (-0.67, -0.40) |
| **Wave 1** | Wave 3 | low | 2.09 (2.63) | 0.00 (ref) |
| | | moderate | 1.71 (2.43) | -0.20 (-0.31, -0.09) |
| | | high | 1.14 (2.01) | -0.66 (-0.80, -0.52) |
| **Wave 2** | Wave 3 | low | 1.79 (2.46) | 0.00 (ref) |
| | | moderate | 1.58 (2.37) | -0.20 (-0.30, -0.10) |
| | | high | 1.46 (2.29) | -0.26 (-0.42, -0.10) |

All analyses were adjusted for age at assessment, sex, race/ethnicity, family annual income, parental average education year, pubertal score at assessment, and waist-to-height ratio at the assessment.
MIND, the mediterranean dietary approaches to stop hypertension intervention for neurodegenerative delay; CBCL, the Child Behavior Checklist.

**Table S6. Cross-sectional and longitudinal associations between adherence to MIND diet and clinical depression additionally adjusting for waist-to-height ratio at the assessment**

| Time of MIND | Time of CBCL | Subgroup of MIND | N (%) | Odds Ratio (95%CI) |
|---|---|---|---|---|
| **Wave 1** | Wave 1 | low | 52 (5.5%) | 0.00 (ref) |
| | | moderate | 48 (2.7%) | 0.54 (0.33,0.88) |
| | | high | 20 (1.9%) | 0.36 (0.19,0.68) |
| **Wave 2** | Wave 2 | low | 55 (3.9%) | 0.00 (ref) |
| | | moderate | 59 (3.1%) | 0.79 (0.54,1.17) |
| | | high | 12 (3.0%) | 0.76 (0.39,1.46) |
| **Wave 3** | Wave 3 | low | 55 (3.7%) | 0.00 (ref) |
| | | moderate | 71 (3.7%) | 0.98 (0.68,1.41) |
| | | high | 2 (0.5%) | 0.13 (0.03,0.53) |
| **Wave 1** | Wave 2 | low | 48 (5.1%) | 0.00 (ref) |
| | | moderate | 51 (2.9%) | 0.59 (0.37,0.93) |
| | | high | 27 (2.6%) | 0.5 (0.29,0.86) |
| **Wave 1** | Wave 3 | low | 38 (4.0%) | 0.00 (ref) |
| | | moderate | 66 (3.8%) | 0.98 (0.65,1.49) |
| | | high | 24 (2.3%) | 0.56 (0.33,0.95) |
| **Wave 2** | Wave 3 | low | 54 (3.8%) | 0.00 (ref) |
| | | moderate | 63 (3.3%) | 0.87 (0.60,1.27) |
| | | high | 11 (2.7%) | 0.72 (0.37,1.41) |

All analyses were adjusted for age at assessment, sex, race/ethnicity, family annual income, parental average education year, pubertal score at assessment, and waist-to-height ratio at the assessment. MIND, the mediterranean dietary approaches to stop hypertension intervention for neurodegenerative delay; PRS, polygenic risk score; CBCL, the Child Behavior Checklist.

**Table S7. Cross-sectional and longitudinal associations between adherence to MIND diet and depression symptom score, stratified based on whether BMI exceeds the 85th percentile**

| Time of MIND | Time of CBCL | Subgroup for BMI | Adjusted β (95%CI ) | | |
|---|---|---|---|---|---|
| | | | Low MIND diet | Moderate MIND diet | High MIND diet |
| **Wave 1** | Wave 1 | BMI p≥85 | 0.00 (ref) | -0.26 (-0.40, -0.13) | -0.73 (-0.90, -0.57) |
| | | BMI p<85 | 0.00 (ref) | -0.46 (-0.55, -0.36) | -0.77 (-0.89, -0.65) |
| **Wave 2** | Wave 2 | BMI p≥85 | 0.00 (ref) | -0.13 (-0.25, -0.01) | -0.21 (-0.40, -0.02) |
| | | BMI p<85 | 0.00 (ref) | -0.18 (-0.27, -0.1) | -0.14 (-0.28, 0.01) |
| **Wave 3** | Wave 3 | BMI p≥85 | 0.00 (ref) | -0.12 (-0.38, 0.13) | -0.36 (-0.79, 0.07) |
| | | BMI p<85 | 0.00 (ref) | -0.06 (-0.25, 0.12) | -0.21 (-0.54, 0.12) |
| **Wave 1** | Wave 2 | BMI p≥85 | 0.00 (ref) | -0.20 (-0.33, -0.06) | -0.57 (-0.73, -0.41) |
| | | BMI p<85 | 0.00 (ref) | -0.33 (-0.43, -0.23) | -0.60 (-0.71, -0.48) |
| **Wave 1** | Wave 3 | BMI p≥85 | 0.00 (ref) | -0.18 (-0.31, -0.05) | -0.69 (-0.85, -0.53) |
| | | BMI p<85 | 0.00 (ref) | -0.29 (-0.39, -0.20) | -0.64 (-0.75, -0.52) |
| **Wave 2** | Wave 3 | BMI p≥85 | 0.00 (ref) | -0.20 (-0.31, -0.08) | -0.23 (-0.42, -0.04) |
| | | BMI p<85 | 0.00 (ref) | -0.13 (-0.22, -0.05) | -0.14 (-0.29, 0.003) |

All analyses were adjusted for age at assessment, sex, race/ethnicity, family annual income, Body Mass Index at assessment, parental average education year, and pubertal score at assessment.

MIND, the mediterranean dietary approaches to stop hypertension intervention for neurodegenerative delay; PRS, polygenic risk score; CBCL, the Child Behavior Checklist.

**Table S8. Cross-sectional and longitudinal associations between adherence to MIND diet and withdraw depression score**

| Time of MIND | Time of CBCL | Subgroup of MIND | Mean (SD) | Adjusted β (95%CI) |
|---|---|---|---|---|
| **Wave 1** | Wave 1 | low | 1.36 (1.97) | 0.00 (ref) |
| | | moderate | 1.05 (1.70) | -0.25 (-0.33, -0.17) |
| | | high | 0.82 (1.46) | -0.51 (-0.61, -0.41) |
| **Wave 2** | Wave 2 | low | 1.29 (2.01) | 0.00 (ref) |
| | | moderate | 1.14 (1.83) | -0.08 (-0.15, -0.01) |
| | | high | 1.14 (1.83) | -0.08 (-0.20, 0.03) |
| **Wave 3** | Wave 3 | low | 1.58 (2.25) | 0.00 (ref) |
| | | moderate | 1.36 (2.06) | -0.12 (-0.19, -0.06) |
| | | high | 1.13 (1.72) | -0.27 (-0.39, -0.16) |
| **Wave 1** | Wave 2 | low | 1.46 (2.07) | 0.00 (ref) |
| | | moderate | 1.20 (1.89) | -0.18 (-0.26, -0.10) |
| | | high | 0.96 (1.71) | -0.41 (-0.51, -0.31) |
| **Wave 1** | Wave 3 | low | 1.80 (2.35) | 0.00 (ref) |
| | | moderate | 1.43 (2.10) | -0.23 (-0.31, -0.16) |
| | | high | 1.10 (1.86) | -0.49 (-0.59, -0.40) |
| **Wave 2** | Wave 3 | low | 1.53 (2.20) | 0.00 (ref) |
| | | moderate | 1.37 (2.08) | -0.10 (-0.17, -0.03) |
| | | high | 1.38 (2.02) | -0.08 (-0.19, 0.03) |

All analyses were adjusted for age at assessment, sex, race/ethnicity, family annual income, Body Mass Index at assessment, parental average education year, and pubertal score at assessment. MIND, the mediterranean dietary approaches to stop hypertension intervention for neurodegenerative delay; CBCL, the Child Behavior Checklist.

**Table S9. Cross-sectional and longitudinal associations between adherence to MIND diet and withdraw depression**

| Time of MIND | Time of CBCL | Subgroup of MIND | N (%) | Odds Ratio (95%CI) |
|---|---|---|---|---|
| **Wave 1** | Wave 1 | low | 83 (3.9%) | 1.00 (ref) |
| | | moderate | 96 (2.4%) | 0.70 (0.51,0.95) |
| | | high | 38 (1.6) | 0.46 (0.31,0.68) |
| **Wave 2** | Wave 2 | low | 84 (2.7%) | 1.00 (ref) |
| | | moderate | 96 (2.1%) | 0.81 (0.59,1.11) |
| | | high | 16 (1.9%) | 0.70 (0.39,1.23) |
| **Wave 3** | Wave 3 | low | 120 (3.4%) | 1.00 (ref) |
| | | moderate | 101 (2.4%) | 0.69 (0.53,0.91) |
| | | high | 12 (1.5%) | 0.43 (0.24,0.78) |
| **Wave 1** | Wave 2 | low | 60 (2.8%) | 1.00 (ref) |
| | | moderate | 92 (2.3%) | 0.87 (0.62,1.21) |
| | | high | 44 (1.9%) | 0.68 (0.46,1.01) |
| **Wave 1** | Wave 3 | low | 81 (3.8%) | 1.00 (ref) |
| | | moderate | 111 (2.8%) | 0.74 (0.55,0.99) |
| | | high | 41 (1.8%) | 0.47 (0.32,0.69) |
| **Wave 2** | Wave 3 | low | 94 (3.0%) | 1.00 (ref) |
| | | moderate | 120 (2.7%) | 0.88 (0.67,1.16) |
| | | high | 19 (2.2%) | 0.72 (0.44,1.19) |

All analyses were adjusted for age at assessment, sex, race/ethnicity, family annual income, Body Mass Index at assessment, parental average education year, and pubertal score at assessment. The age- and sex-adjusted T-score no less than 70 was corresponding to higher than the $98^{th}$ percentile in the general population, thus defined as withdraw depression.

MIND, the mediterranean dietary approaches to stop hypertension intervention for neurodegenerative delay; PRS, polygenic risk score; CBCL, the Child Behavior Checklist.

**Table S10. Longitudinal associations between adherence to MIND diet and depression symptom score, additionally adjusting for the depression symptom score at the assessment for MIND diet**

| Time of MIND | Time of CBCL | Subgroup of MIND | Mean (SD) | Adjusted β (95%CI) |
|---|---|---|---|---|
| **Wave 1** | Wave 2 | low | 1.88 (2.48) | 0.00 (ref) |
| | | moderate | 1.45 (2.22) | -0.10 (-0.16, -0.03) |
| | | high | 1.07 (1.92) | -0.27 (-0.35, -0.19) |
| **Wave 1** | Wave 3 | low | 2.12 (2.70) | 0.00 (ref) |
| | | moderate | 1.69 (2.51) | -0.07 (-0.14, -0.004) |
| | | high | 1.21 (2.31) | -0.35 (-0.43, -0.27) |
| **Wave 2** | Wave 3 | low | 1.81 (2.58) | 0.00 (ref) |
| | | moderate | 1.58 (2.43) | -0.08 (-0.14, -0.03) |
| | | high | 1.54 (2.40) | -0.11 (-0.20, -0.01) |

All analyses were adjusted for age at assessment, sex, race/ethnicity, family annual income, Body Mass Index at assessment, parental average education year, pubertal score at assessment, and the depression symptom score at the assessment for MIND diet.

MIND, the mediterranean dietary approaches to stop hypertension intervention for neurodegenerative delay; PRS, polygenic risk score; CBCL, the Child Behavior Checklist.

**Table S11. The predictive analysis between MIND and clinical depression additionally adjusting for whether clinical depression at the time for MIND diet**

| Time of MIND | Time of CBCL | Subgroup of MIND | N (%) | Odds Ratio (95%CI) |
|---|---|---|---|---|
| **Wave 1** | Wave 2 | low | 95 (4.5%) | 1.00 (ref) |
| | | moderate | 120 (3.0%) | 0.85 (0.62, 1.17) |
| | | high | 58 (2.5%) | 0.77 (0.52, 1.12) |
| **Wave 1** | Wave 3 | low | 104 (4.9%) | 1.00 (ref) |
| | | moderate | 151 (3.8%) | 0.95 (0.71, 1.25) |
| | | high | 62 (2.7%) | 0.70 (0.49, 0.98) |
| **Wave 2** | Wave 3 | low | 131 (4.2%) | 1.00 (ref) |
| | | moderate | 161 (3.6%) | 0.92 (0.71, 1.19) |
| | | high | 25 (2.9%) | 0.77 (0.49, 1.23) |

All analyses were adjusted for age at assessment, sex, race/ethnicity, family annual income, Body Mass Index at assessment, parental average education year, pubertal score at assessment, and the depression symptom score at the assessment for MIND diet.

MIND, the mediterranean dietary approaches to stop hypertension intervention for neurodegenerative delay; PRS, polygenic risk score; CBCL, the Child Behavior Checklist.

**Table S11. CLPM model fit indices across waves**

| | **cbcl_scr_dsm5_depress_r** |
|---|---|
| **Test statistic** | 638.501 |
| **Degrees of freedom** | 4 |
| **P-value (Chi-square)** | 0 |
| **Robust Comparative Fit Index (CFI)** | 0.932 |
| **Robust Tucker-Lewis Index (TLI)** | 0.743 |
| **Robust RMSEA** | 0.137 |
| **SRMR** | 0.036 |

CLPM, cross-lagged panel model; MIND, the mediterranean dietary approaches to stop hypertension intervention for neurodegenerative delay; RMSEA, root mean square error of approximation; SRMR, standardized root mean square residual.

**Table S12. Cross-sectional and longitudinal associations between adherence to MIND diet and depression symptom score additionally adjusting for PRS of depression symptoms.**

| **Time of MIND** | **Time of CBCL** | **Subgroup for PRS** | **Adjusted β (95%CI )** | | |
|---|---|---|---|---|---|
| | | | Low MIND diet | Moderate MIND diet | High MIND diet |
| **Wave 1** | Wave 1 | Low Risk | 0.00 (ref) | -0.4 (-0.62, -0.18) | -0.72 (-1.01, -0.44) |
| | | Moderate Risk | 0.00 (ref) | -0.43 (-0.63, -0.23) | -0.71 (-0.95, -0.47) |
| | | Severe Risk | 0.00 (ref) | -0.44 (-0.63, -0.25) | -0.84 (-1.09, -0.59) |
| **Wave 2** | Wave 2 | Low Risk | 0.00 (ref) | -0.18 (-0.37, 0.01) | 0.21 (-0.12, 0.54) |
| | | Moderate Risk | 0.00 (ref) | -0.16 (-0.36, 0.03) | -0.36 (-0.67, -0.04) |
| | | Severe Risk | 0.00 (ref) | -0.14 (-0.31, 0.04) | -0.35 (-0.67, -0.03) |
| **Wave 3** | Wave 3 | Low Risk | 0.00 (ref) | -0.38 (-0.57, -0.19) | -0.5 (-0.84, -0.17) |
| | | Moderate Risk | 0.00 (ref) | -0.11 (-0.30, 0.07) | -0.19 (-0.5, 0.12) |
| | | Severe Risk | 0.00 (ref) | -0.08 (-0.25, 0.09) | -0.32 (-0.64, 0.00) |
| **Wave 1** | Wave 2 | Low Risk | 0.00 (ref) | -0.27 (-0.47, -0.06) | -0.65 (-0.92, -0.38) |
| | | Moderate Risk | 0.00 (ref) | -0.37 (-0.58, -0.16) | -0.6 (-0.86, -0.35) |
| | | Severe Risk | 0.00 (ref) | -0.37 (-0.56, -0.18) | -0.64 (-0.88, -0.4) |
| **Wave 1** | Wave 3 | Low Risk | 0.00 (ref) | -0.29 (-0.5, -0.09) | -0.7 (-0.97, -0.44) |
| | | Moderate Risk | 0.00 (ref) | -0.23 (-0.44, -0.02) | -0.46 (-0.71, -0.21) |
| | | Severe Risk | 0.00 (ref) | -0.39 (-0.57, -0.21) | -0.76 (-1.00, -0.53) |
| **Wave 2** | Wave 3 | Low Risk | 0.00 (ref) | -0.15 (-0.34, 0.05) | -0.24 (-0.59, 0.11) |
| | | Moderate Risk | 0.00 (ref) | -0.08 (-0.27, 0.12) | -0.13 (-0.43, 0.17) |
| | | Severe Risk | 0.00 (ref) | -0.11 (-0.28, 0.07) | -0.42 (-0.73, -0.11) |

All analyses were adjusted for age at assessment, sex, race/ethnicity, family annual income, Body Mass Index at assessment, parental average education year, and pubertal score at assessment.
MIND, the mediterranean dietary approaches to stop hypertension intervention for neurodegenerative delay; PRS, polygenic risk score; CBCL, the Child Behavior Checklist.

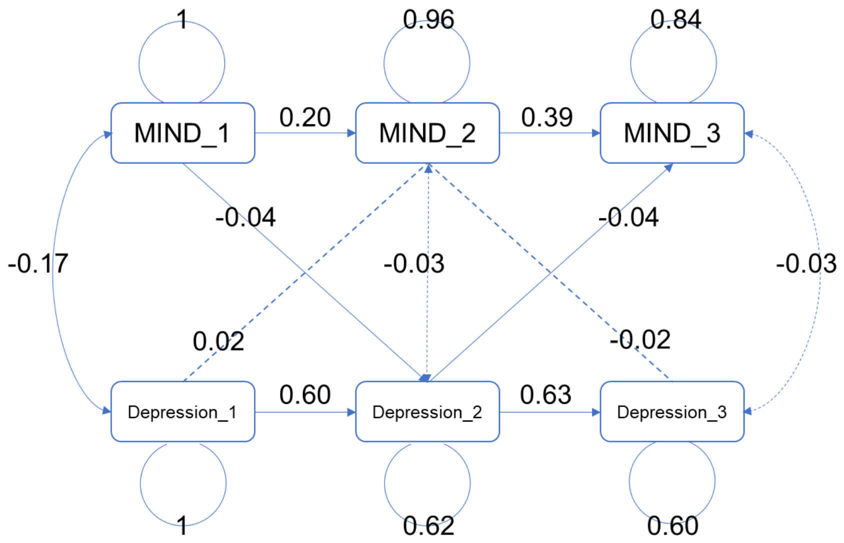

**Figure S1. CLPM of MIND and depression symptom score additionally adjusting for PRS.**

All analyses were adjusted for age at assessment, sex, race/ethnicity, family annual income, Body Mass Index at assessment, parental average education year, pubertal score at assessment and PRS of depression symptoms.

CLPM, cross-lagged panel model; MIND, the mediterranean dietary approaches to stop hypertension intervention for neurodegenerative delay; PRS, polygenic risk score.
